# Anterior Talofibular Ligament and Calcaneofibular Ligament Repair Reinforced by Bio-inductive Implants

**DOI:** 10.1101/2025.09.11.25335447

**Authors:** Atta Taseh, Lercan Aslan, Mohammad Hossein Chegini Kord, Mohamad Al Masri, Ahmad Hedayatzadeh Razavi, Nazanin Nafisi, Colin O’Neill, Kendal Toy, Joaquin Batista, Gregory Waryasz, Christopher P. Miller, Ara Nazarian, Soheil Ashkani-Esfahani

**Author notes:** **Corresponding author:**. Atta Taseh, Address: 158 Boston Post Road, Weston, MA, 02493. Co-first authors (Atta Taseh, Lercan Aslan). Co-senior authors (Ara Nazarian, Soheil Ashkani-Esfahani).

## Abstract

**Background:** Chronic lateral ankle instability stems from laxity in the anterior talofibular (ATFL) and calcaneofibular (CFL) ligaments. While suture repair is standard, added reinforcement is often needed to restore native strength. This study evaluates the biomechanics bio-inductive implant augmentation (Bio-Aug) of suture repair.

**Methods:** Eight fresh-frozen human ankle specimens were tested under three ATFL and CFL conditions, including: (1) Intact, (2) Suture-Repair, and (3) Bio-Aug, (BioBrace®, CONMED Co, Largo, FL). Biomechanical testing included a 500-cycle loading protocol, with creep, primary stiffness, and final stiffness as outcomes.

**Results:** Only 32% of Suture-Repair specimens completed all 500 cycles, compared to full completion in other groups (*P* = .04). Bio-Aug results were comparable to Intact, while Suture-Repair showed lower final stiffness than Bio-Aug (*P* = .049). The Intact group had the least creep, although Suture-Repair showed high variability.

**Conclusion:** Bio-inductive implant appears to provide comparable stiffness to native ligaments by reinforcing suture repair.

## 1 INTRODUCTION

Ankle sprains are common musculoskeletal injuries, with over 3 million cases annually. Although most resolve with nonsurgical treatment, about 20% progress to chronic lateral ankle instability (CLAI).[1], [2] The injury mechanism involves forced plantarflexion and inversion, placing excessive stress on the anterior talofibular ligament (ATFL) and calcaneofibular ligament (CFL), primary stabilizers of the lateral ankle. Disruption of these ligaments is closely linked to the development of CLAI and may require surgical intervention if conservative treatment fails.[3]

Historically, several surgical approaches have been proposed for the treatment of CLAI, with the Broström procedure and its subsequent modifications being among the most commonly performed.[4] While this approach has demonstrated favorable clinical outcomes, some patients exhibit residual instability postoperatively, particularly in cases with ligamentous attenuation or poor tissue quality.[5] Furthermore, the procedure requires a long post-operative immobilization period for optimal tissue healing.[6] In recent years, alternative techniques such as suture anchor fixation, suture tape augmentation, anatomic reconstruction using autografts or allografts, and arthroscopic techniques have gained traction.[7] These approaches aim to provide enhanced early stabilization and optimal support for tissue healing, potentially mitigating some of the limitations associated with the traditional Broström repair.

Bio-inductive implants have emerged as a promising adjunct to traditional ligament repair. These implants, composed of either collagen-based or synthetic polymeric matrices, are designed to enhance biological healing and augment structural integrity by promoting tissue formation.[8], [9] Recent advancements in collagen-based polymeric bio-composites have demonstrated enhanced mechanical strength and structural stability, facilitating the gradual development of organized fibrous tissue essential for successful tissue reconstruction or engineering.[10], [11], [12] The efficacy of bio-inductive implants was reported in rotator cuff repairs, anterior cruciate ligament reconstructions, and Achilles tendon repair.[13] Given the different mechanisms of injury in CLAI and the biomechanical importance of precise ligamentous tensioning in restoring ankle stability, it remains unclear whether augmentation with bio-inductive implants can provide superior mechanical properties compared to traditional repair techniques.[14]

In this study, we aimed to evaluate the mechanical properties of ATFL and CFL ligament repair using a composite bio-inductive implant comprised of a highly porous type I collagen matrix reinforced with bioresorbable poly-L-lactic acid (PLLA) microfilaments. We assessed stiffness and creep behavior under cyclic loading conditions to determine whether bio-inductive implant augmentation offers a mechanical advantage over suture repair.

## 2 MATERIALS AND METHODS

### 2.1 Study Design and Specimens

Eight fresh-frozen lower extremity knee-disarticulated human specimens. The mean age of the specimens was 60 years old with 54 as the youngest and 76 as the oldest. Cadavers with evidence of injury upon surgical exploration were excluded from the study. All cadavers were preserved at –20°C and thawed at room temperature prior to use.

All specimens were tested under three different testing conditions, including (1) Intact ligaments group, (2) Suture-Repair group (ruptured and repaired with suture), and (3) Bio-inductive Implant Augmentation (Bio-Aug) group (ruptured and repaired with BioBrace®, CONMED Co, Largo, FL). Suture repair or repair plus augmentation was performed randomly to minimize the bias due to the effect of soft tissue changes in each testing condition. Surgical procedures were performed by four experienced orthopaedic surgeons following the established study protocol for consistency.[14]

### 2.2 Surgical procedure

The procedure began with preparing the specimens for intact testing. An incision was made around the ankle joint, including a curvilinear incision on the lateral side, approximately 3 cm distal to the tip of the fibula. All lower leg muscles, including the extrinsic foot muscles, gastrocnemius, and soleus, along with the associated skin, were excised. Tibial ligamentous attachments on the anterior, medial, and posterior sides were resected. The proximal portion of the interosseous membrane was resected along with ligaments, including the anterior inferior tibiofibular ligament (AITFL), posterior inferior tibiofibular ligament (PITFL), distal interosseous membrane, and transverse tibiofibular ligament (TTFL). Finally, after excising the tibia, the posterior talofibular ligament and the peroneal retinaculum were resected while making sure the CFL, ATFL, and joint capsule were intact. The anterior and posterior borders of the fibula were marked on the talar cartilage and used as the reference position during the subsequent testing conditions. The ATFL and CFL were identified and marked at their origin and insertion sites to facilitate identification during the repair. Both ligaments were resected mid-substance from the medial side to preserve the integrity of the joint capsule, which was retained for testing (Figure 1).

**Figure 1.**
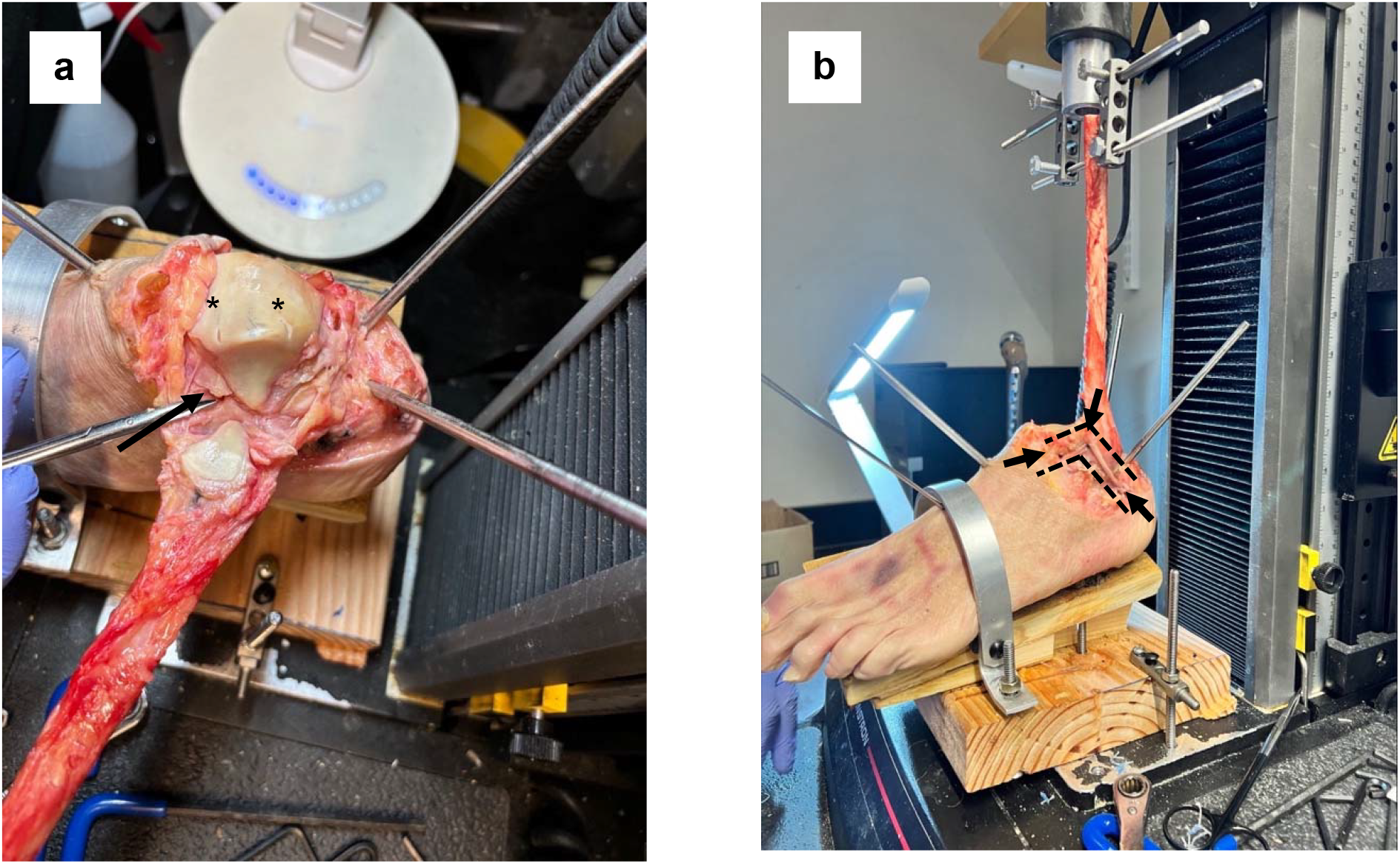
Experimental setup demonstrating (a) Talar reference markings (asterisks) utilized for fibular alignment, with an arrow indicating the resected anterior talofibular ligament (ATFL); (b) Proper positioning of the foot stabilized by a strapped jig and Steinmann pins, illustrating axial force application through the fibula. The dashed line outlines the triangular trajectory of the bio-inductive implant, with arrows marking the insertion points.

To perform the suture repair, the fibula was realigned to its pre-determined reference position on the talar cartilage during the procedure (Figure 1). The anatomical origins and insertions of the CFL and ATFL were precisely identified based on previous markings. Both ligaments were repaired using a braided, non-absorbable suture (Hi-Fi®, CONMED Co, Largo, FL) and a continuous suturing technique, which involved two passes to ensure secure reattachment and achieve optimal tension. Additionally, the joint capsule between the ATFL and CFL, as well as the capsule posterior to the CFL, was repaired with two suture passes per site to restore integrity and support.

To perform augmentation, the bio-inductive implant was fashioned into a triangular configuration, with its apex anchored to the proximal one-third of the anterior border of the lateral malleolus and its remaining vertices positioned at the talar neck and calcaneus, aligning with the anatomical course of the ATFL and CFL (Figure 1). To secure the implant, 2.4 mm anchor holes were created at the designated anatomical regions using guide pins and cannulated drills to ensure precise and controlled placement. Suture anchors (Argo Knotless®, CONMED Co, Largo, FL) were then employed to fix the implant. Each anchor was loaded with the implant material such that 2 mm of the implant passed through the anchor eyelet. Anchors were subsequently inserted flush with the cortical bone surface to achieve uniform tensioning across all specimens. This approach was designed to provide optimal mechanical support while maintaining anatomical fidelity.

### 2.3 Mechanical Testing and Study Outcomes

The specimens were mounted onto a mechanical testing machine (Instron 5944, Instron, Norwood, MA) using a custom-designed jig (Figure 1). This setup positioned the foot at 20° of inversion and 10° of plantar flexion, simulating the tension experienced by the ATFL and CFL, as described by Waldrop et al.[14]The jig incorporated an adjustable metal strap across the midfoot to accommodate varying cadaver sizes and secure the specimen. To prevent sliding or rotational movements, two Steinmann pins were inserted through the subtalar joint and two in the midfoot area (Figure 1). Additionally, a 5-millimeter Steinmann pin was inserted into the proximal part of the fibula to connect the specimen to the mechanical testing machine. The alignment of the fibula was carefully adjusted to ensure it was vertically aligned with the actuator axis, and the entire assembly was fixed to the base of the mechanical testing device.

All specimens underwent a pre-conditioning protocol of 20 cycles between 10 N and 20 N at a loading rate of 5 N/s prior to testing. The cyclic loading consisted of 500 cycles ranging from 20 N (minimum) to 100 N (peak force), applied at a loading rate of 50 N/s. No designated rest periods occurred between cycles. However, a standardized 20-minute rest period was incorporated between testing conditions to reduce fatigue effects and simulate physiological recovery. Following cyclic loading, specimens in the Bio-Aug group were subjected to load-to-failure testing at a force application rate of 20 N/s. To prevent tissue desiccation, specimens were consistently sprayed with phosphate-buffered saline (PBS) every 100 cycles during the cyclic loading phase. Hydration was also maintained during setup and in the intervals between tests by reapplying PBS solution regularly to the exposed tissues. Testing was performed at the room-temperature (approximately 22–23°C).

Force-displacement data were collected using the mechanical testing machine to evaluate the biomechanical properties of the ATFL and CFL. The data consisted of cyclic loading profiles with displacement and force recorded at high resolution (Figure 2). From this dataset, two key metrics, creep and stiffness, were calculated to assess the viscoelastic behavior and structural integrity of the ligaments. Furthermore, load-to-failure was tested for the Bio-Aug group upon completion of the cyclic loading testing conditions.

**Figure 2.**
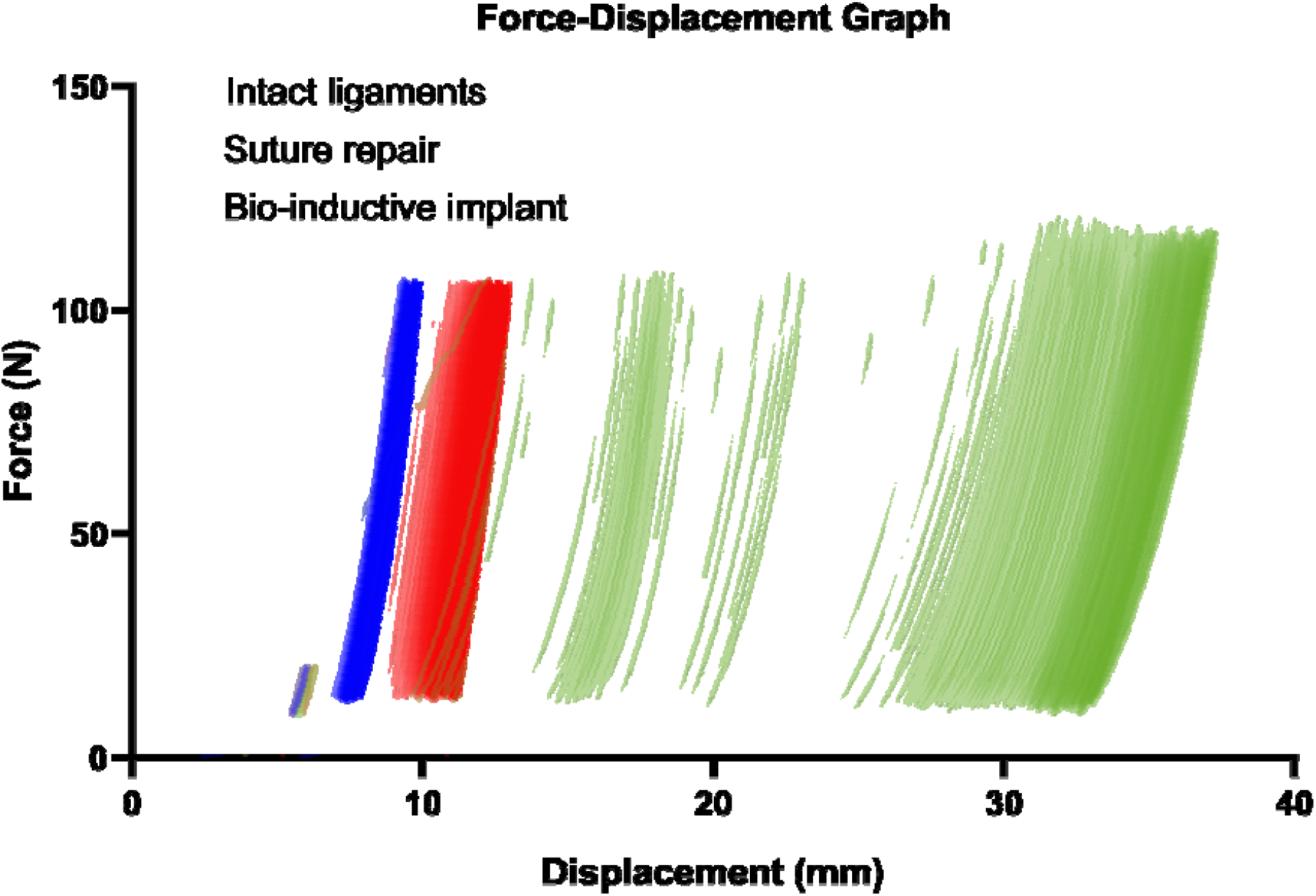
Force-Displacement graph of a single specimen examined under three different conditions.

Creep is defined as a viscoelastic response of tissues under sustained loading and comprises three distinct phases: primary, secondary, and tertiary creep.[15] These phases describe how ligament displacement evolves under constant cyclic loading. The rate of deformation decreases as the fibers resist further elongation and begin to stabilize. Primary creep is characterized by rapid initial deformation while secondary creep happens following the primary phase when the deformation slows down and enters a steady-state phase. In this study, we focused on primary creep due to early specimen failure preventing progression to secondary creep (Figure 3). Primary creep was quantified by fitting an exponential function (y=a.e^(b.x)+c; where a, b, c are called the parameters) to the displacement data from the force-displacement curve (Figure 3). This model allowed us to identify the equilibrium point where deformation plateaued, marking the transition to secondary creep.

**Figure 3.**
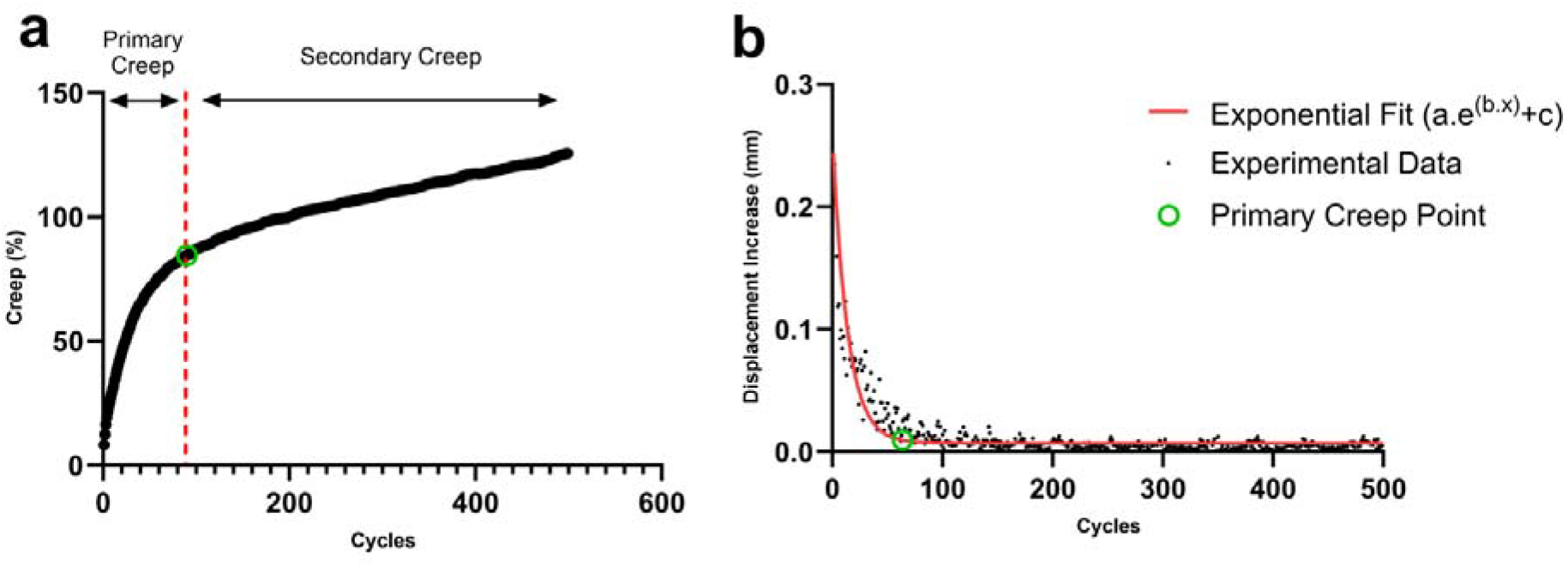
Creep Analysis: (a) Creep percentage over cycles, highlighting primary and secondary creep phases. (b) Displacement over cycles with an exponential fit used to quantify primary creep

Stiffness, defined as the slope of the force-displacement relationship during loading, was calculated for three key cycles: the primary creep cycle and the final cycle. Cycles were identified by detecting force increases that crossed a defined threshold of 100 N, and the data for each cycle’s loading phase were extracted. Linear regression was applied to the loading segments of the force-displacement data for these specific cycles to calculate stiffness as the slope (ΔF/ΔD).[16] This approach ensured that stiffness was assessed at representative intervals, capturing the ligament’s structural response during primary creep and by the end of the cyclic test.

### 2.4 Statistical Analysis

Statistical analysis was conducted using SPSS v28.0 (SPSS, Chicago, IL). Data distribution was assessed with the Shapiro-Wilk test. Creep, stiffness, and the percentage of completed cycles were compared across groups using the Wilcoxon Signed-Rank Test, with a P-value < .05 indicating statistical significance.

## 3 RESULTS

Eight specimens were included in the study. All specimens in both the Intact and Bio-Aug groups successfully completed the full 500-cycle mechanical loading protocol. In contrast, the Suture-Repair group showed lower resilience, with only three of eight specimens (37.5%) completing the full loading protocol (Table 1). This difference in survivability was statistically significant (*P* = .04).

**Table 1.** Summary of survival and failure data of study specimens, with all groups subjected to cyclic loading tests up to 500 cycles.

| Testing Condition | Total number of specimens | Survived all 500 cycles | Percentage of cycles completed* |
| --- | --- | --- | --- |
| Intact ligaments | 8 | 8 | 100 (100,100) |
| Suture repair | 8 | 3 | 32** (1.5, 100) |
| Bio-inductive implant repair | 8 | 8 | 100 (100, 100) |

\* Median (IQR) values are presented
\*\* Significantly different from other groups, $P=.04$

Stiffness measurements, taken at both the beginning and end of the cyclic loading protocol, further highlighted mechanical differences across the three groups. Primary stiffness was highest in the Intact group, followed closely by the Bio-Aug group, and lowest in the Suture-Repair group. The Suture-Repair group exhibited significantly lower stiffness than the Intact group (*P* = .049) and trended lower than the Bio-Aug group, although this difference did not reach statistical significance (Table 2). Final stiffness, assessed at the 500th cycle or the final completed cycle for failed specimens, followed a similar pattern. The Suture-Repair group demonstrated the lowest stiffness, with values significantly lower than the Bio-Aug group (*P* = .049).

**Table 2.** A comparison of stiffness data [median (IQR)] at primary, and final time points for study groups under cyclic loading testing.

| Testing condition | Primary stiffness (N/mm) | Final stiffness (N/mm) |
| --- | --- | --- |
| Intact ligaments | 38.37 (25.12, 43.09) | 41.72 (27.32, 51.31) |
| Suture repair | 27.6 (5.9, 43) | 30.31 (6.22, 44.18) |
| Bio-inductive<br>implant<br>augmentation | 38.64 (30.54, 45.29) | 43.26 (33.24, 52.52) |
| *P*-value * | .049 **, .26<sup>†</sup>, .07<sup>‡</sup> | .07 **, .4<sup>†</sup>, .049<sup>‡</sup> |

\* Wilcoxon Signed-Rank Test
<sup>†</sup> Intact ligaments vs Bio-inductive implant repair
\*\* Intact ligaments vs Suture repair
<sup>‡</sup> Bio-inductive implant repair vs Suture repair

The viscoelastic behavior of the constructs, as characterized by primary creep, also varied among groups. The Suture-Repair group displayed the greatest creep, reflecting increased laxity and diminished ability to resist deformation. In contrast, the Intact group had the lowest creep, while the Bio-Aug group showed intermediate values (Figure 4).

**Figure 4.**
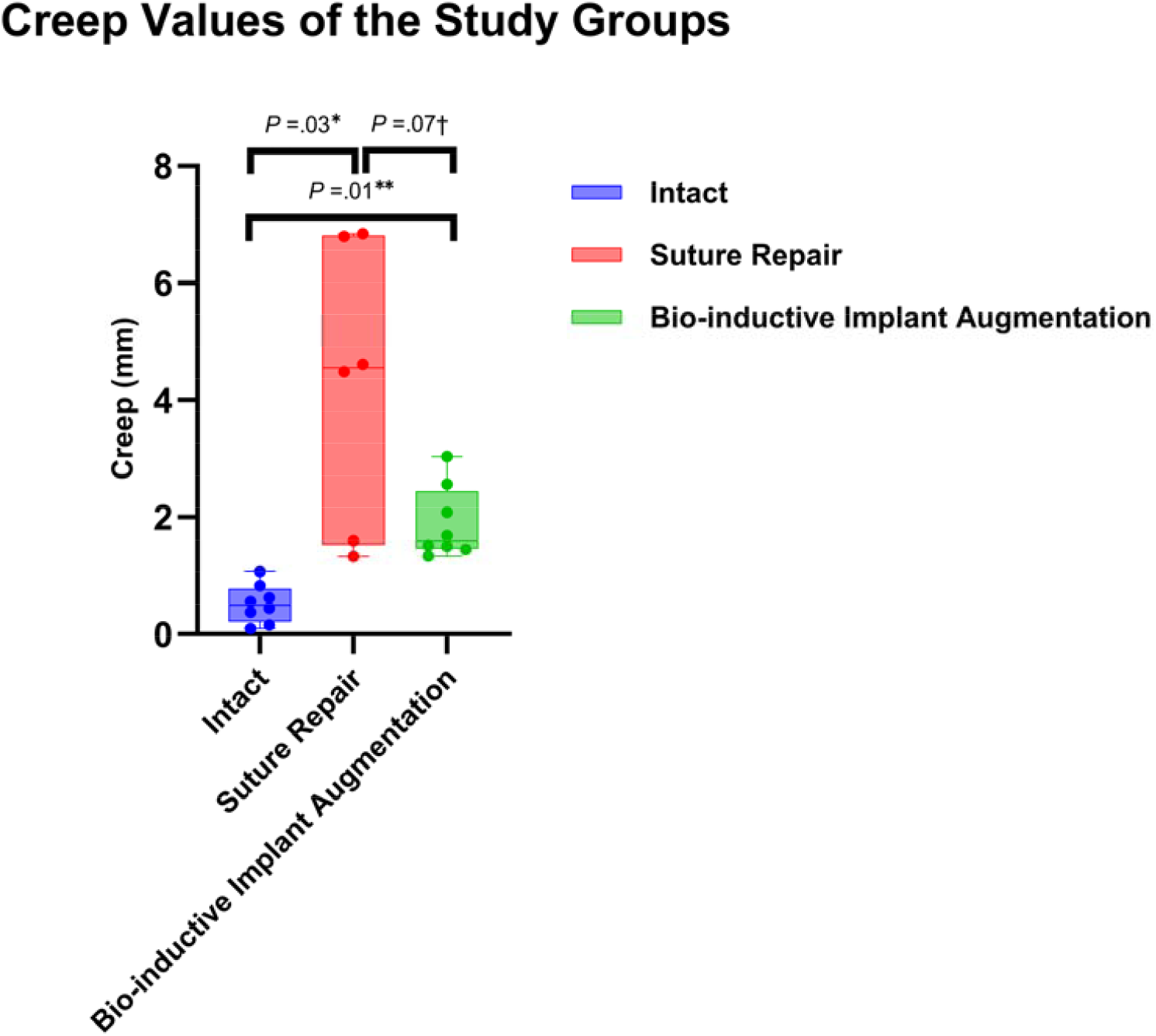
Median values of the creep measurements across the three experimental groups. Bars represent the interquartile ranges within each group, and the *P*-values highlights the statistical difference between the groups (*Intact vs Suture-Repair, **Intact vs Bio-inductive Implant Augmentation, †Suture-Repair vs Bio-inductive Implant Augmentation).

Finally, to evaluate post-cycling structural strength, four Bio-Aug specimens were subjected to load-to-failure testing. The median failure load was 330 N (IQR: 300–360 N). Due to the randomized experimental design, post-cycling load-to-failure testing was intended for four specimens in each of the Bio-Aug and Suture-Repair groups. However, all four Suture-Repair specimens failed during cyclic loading, preventing subsequent load-to-failure testing in that group.

## 4 DISCUSSION

Our study evaluated the biomechanical effectiveness of a bio-inductive implant for augmentation after ATFL and CFL repair, with and without concomitant suture repair. The results demonstrated that this implant successfully restored ligament stiffness and resistance to cyclic loading, achieving values approaching those of the intact ligament. These findings suggest that bio-inductive implants may serve as an effective augmentation material, providing more durable and near-physiological ligament properties, thereby potentially reducing the need for extended immobilization periods following standard repair.

The modified Broström-Gould procedure is the most common treatment for CLAI[17]. However, it necessitates extended immobilization periods to prevent ligament elongation and provides limited initial mechanical strength, which can delay accelerated physical therapy, which is particularly problematic for athletes.[18], [19] The introduction of augmentation strategies has addressed these shortcomings by offering increased biomechanical strength, facilitating earlier and more intensive rehabilitation protocols, thus yielding improved functional outcomes.[18], [20], [21], [22], [23] Highlighting these limitations, Maffulli et al. reported that only 58% of athletes returned to their pre-injury level of sports participation following isolated Broström repairs, underscoring the inadequacy of suture repair alone in high-demand patients.[24]

Clinically, achieving near-native stiffness is essential to prevent excessive ligament elongation under repetitive loading during the early phases of recovery, especially in high-demand patients.[14], [25] By matching or slightly exceeding the stiffness of native ligaments, bio-inductive implants may provide sufficient mechanical reinforcement during the early physical therapy. Constructs such as the internal brace, which exhibit significantly higher stiffness compared to native ligaments, may restrict ankle and subtalar range of motion.[14], [25], [26] This concern is supported by Coetzee et al., who demonstrated that patients treated with internal brace augmentation experienced significantly reduced ankle dorsiflexion compared to their non-operated side. ^5^Creep, the progressive elongation of a ligament under sustained cyclic load, is another critical factor influencing repair durability. Furthermore, our results showed significantly lower creep in the Bio-Aug group than in Suture-Repair. Though still higher than the intact ligaments, it is another critical factor enhancing repair durability compared to standard suture repair. These findings are consistent with prior work by Giza et al., who demonstrated that Achilles tendon repairs augmented with a collagen scaffold exhibited reduced elongation under repetitive loading.[27]

Hunt et al., in their review of open Broström techniques, recommended two weeks of non-weight-bearing and a ten-week limitation on the range of motion, especially inversion, to avoid damaging the surgical repair.[28] Conversely, Coetzee et al. demonstrated that patients treated with internal brace augmentation for CLAI could initiate weight-bearing immediately postoperatively, with unrestricted range of motion exercises permitted from six weeks onward.[21] Similarly, Cho et al. reported excellent outcomes without elongation complications in a population with generalized ligamentous laxity after reconstruction with internal brace augmentation.[20] These accelerated rehabilitation protocols are further supported by biomechanical studies indicating that augmented repairs exceed native ligament load-to-failure values by more than 50%, a threshold widely accepted for safely initiating early mobilization.[14], [25] Waldrop et al. reported that isolated Broström repairs resulted in ligament stiffness similar to native tissue but exhibited approximately 42% less ultimate load-to-failure.[14] Consequently, they advised against placing early stress on the repair site, effectively delaying rehabilitation. In contrast, Viens et al. found that the internal brace construct demonstrated an ultimate load-to-failure double that of the native ATFL.[25] While our study did not directly measure the ultimate load-to-failure of native ATFL, previous literature reports consistent values: 154.0 ± 63.7 N by Viens et al., 138.9 ± 23.5 N by Attarian et al., and 160.9 ± 72.2 N by Waldrop et al. Given that the bio-inductive augmentation demonstrated a load-to-failure of approximately 330 N, more than 100% higher than reported values for native ATFL, we suggest that bio-inductive implants could similarly support accelerated physical therapy, comparable to internal brace augmentation.[14], [25], [29]

This study has several limitations that should be acknowledged. First, this time-zero cadaveric study does not account for *in vivo* biological healing or remodeling. Second, the small sample size may limit generalizability. Third, although the extensive soft tissue dissection helped isolate the mechanical contribution of the ATFL and CFL, it created a non-physiological setup that limits the generalizability of our findings to in vivo conditions. Lastly, we standardized the testing order and reserved failure testing for the final stage, however, repeated use of the same specimen may have affected tissue properties.

## 5 CONCLUSION

In conclusion, our study showed that the biomechanical properties of bio-inductive implants, such as stiffness, are similar to those of native ligaments. This distinguishes them from similar constructs with higher stiffness, which may lead to a restricted range of motion. Additionally, the sufficient load-to-failure strength of bio-inductive implants may make them a reliable construct for early physical therapy, which is essential for high-demand patients. Future studies should investigate the clinical performance of bio-inductive implants *in vivo*, including patient-reported outcomes, functional recovery, and long-term durability across different patient populations.

## Acknowledgment

The authors would like to express their deepest gratitude to the individuals who generously donated their bodies to medical research.

## Ethical considerations

The study protocol was approved by the Massachusetts General Hospital Institutional Review Board (IRB # 2016P001295)

## Declaration of Generative AI and AI-assisted technologies in the writing process

During the preparation of this work the authors used OpenAI’s ChatGPT in order to enhance the readability and clarity of this document After using this tool/service, the authors reviewed and edited the content as needed and take(s) full responsibility for the content of the publication

## Data availability

Research data will be made available upon reasonable request, contingent on Institutional Review Board (IRB) approval.

